# The Association Between Threatened Miscarriage And Development Of Gestational Hypertension/Pre-Eclampsia

**DOI:** 10.1101/2021.05.07.21256696

**Authors:** S.M.S.G. Gunarathna, Naleen Ratnayake, Lakshman Pallemulla, D.P. Lanka Rasanjana, P.K. Abeysundara, A.A. Nilanga Nishad

**Affiliations:** Castle Street Hospital for Women, Colombo, Sri Lanka; Worthing Hospital (NHS), Western Sussex, United Kingdom; Center for Epidemiological and Clinical Research LK, Sri Lanka

## Abstract

**Introduction:** Gestational hypertension (GH)/Pre-eclampsia (PEC) is an important cause of direct maternal deaths in Sri Lanka. GH/PEC and threatened miscarriage (TM) share common pathophysiological mechanisms. This study was conducted to determine the association between TM and development of GH/PEC.

**Methodology:** A case control study was conducted at Castle Street Hospital for Women, Sri Lanka from April 2015 to October 2015. Cases consisted of patients with GH/PEC and compared with age and parity matched controls. A systematic random sampling method was used. Similar number of cases and controls were compared while each group consisted of 245 subjects. Data was obtained from medical records. It’s also important to note that mothers aged 20-35 years were included and medical disorders other than GH/PEC was excluded.

**Results:** There were 245 subjects in each group of the study. Among the cases, 56% had GH and the rest had PEC. There were 25 patients with TM in the study population and 64% of them subsequently developed GH or PEC. There is also a significant risk of developing PEC in a patient who had a history of threatened miscarriage (OR 3.31, 95% CI 1.35-8.11). Moreover the patients who had a history of TM tend to develop GH or PEC early, within 20-32 weeks of gestation (OR 11.49, 95% CI 3.88-33.99). As we identified, 62% of patients who had TM developed GH/PEC early (from 20 to 32 weeks) but among the cases who had no history of TM, only 12% developed GH/PEC between 20 to 32 weeks of gestation (O.R. 20.7 (5.66 to 91.96). There is a significant risk of developing severe GH/PEC in the group of patients who had a history of TM (OR 8.59, 95% CI 2.87-25.66). Eighty one percent (81%) of the cases, who had a history of TM, developed severe and moderate GH/PEC rather than mild. But the majority (63%) of the cases, who had no history of TM, developed mild GH/PEC (O.R. 7.6 (2.00 to 42.55).

**Conclusions:** Shared pathophysiological mechanisms of GH/PEC and TM may explain the observed association between these obstetric complications. Early onset, severe GH/PEC in cases with TM suggests temporality and a biological gradient which favors causality.

## Introduction

Hypertension in pregnancy is an important cause of direct maternal deaths in Sri Lanka. Early identification and aggressive treatment of its complications is vital to reduce subsequent morbidity and mortality.(1) Gestational hypertension (GH) is a common medical disorder in pregnancy. Prevalence of pre-eclampsia (PEC) has inflated by nearly one-third throughout the last decade. In addition, hypertensive disorders of pregnancy are accountable for a significant proportion of maternal and perinatal morbidity. It is the second leading cause of maternal mortality globally.(2) Following the global trend, a similar circumstance is seen in Sri Lanka.(3) GH is defined as hypertension detected after 20 weeks of gestation in which chronic hypertension is excluded. When the presence of hypertension along with proteinuria/other laboratory and clinical parameters after 20 weeks is called pre-eclampsia (PEC). Significant proteinuria is only seen in PEC and EC, while occurrence of seizures signifies EC.(4) Chronic hypertension is estimated to affect 2-5% of pregnant women, and is frequently diagnosed during antenatal period. The prevalence of PEC differs with the definition used and the population studied; nevertheless, PEC occurs in less than 5 per cent of an average antenatal population. Hypertensive disorders during pregnancy leads to several maternal complications including eclampsia, HELLP syndrome, cerebral haemorrhage, liver rupture, cortical blindness, pulmonary oedema, placental abruption, cortical blindness and renal failure. Furthermore, intra-uterine growth restriction, preterm delivery and still births are observed adversities to the fetus.(5) Pathophysiological mechanisms responsible for the development of GH/PEC have not yet been fully discovered. Defective utero-placental circulation as a result of sub-optimal trophoblastic invasion of spiral arteries could be the initiating event. This leads to activation of placental endothelium to produce vasoconstrictors (endothelin and thromboxane A2) and suppress production of vasodilators (nitric oxide and prostacyclin).(6) Low dose aspirin (prophylaxis) which inhibits synthesis of thromboxane A2 shows 15% reduction in incidence of PEC. Risk predictors for PEC enable early detection and treatment with aspirin. Aspirin prophylaxis is indicated in high risk pregnancies.

Pregnancies complicated with one or more of past history GH/PEC, chronic hypertension, chronic kidney disease, immune mediated diseases like SLE or APLS and diabetes mellitus are treated with aspirin. Presence of either two or more of the following factors including: an age greater than 40 years, a pregnancy interval of more than 10 years, multiple pregnancies, body mass index of 35kg/m2 and family history of preeclampsia are indicators to treat with aspirin prophylaxis. (7) Threatened miscarriage(TM) was evaluated as an independent risk factor for GH/PEC in previous studies which showed conflicting results.(8) One quarter of all pregnancies is complicated with TM and most of them continue till term, but some of them may progress to missed, incomplete or complete miscarriages.

Limited number of studies have been carried out in Sri Lanka to assess the effects of TM on adverse maternal outcomes.(9) However, none of them specifically focused on association of TM and GH/PEC. Therefore we wanted to determine the association between TM and the development of GH/PEC.

## Methods

This study was carried out as a case control study at ward 3, CSHW for 6 months period from 1^st^ of April 2015 to 1^st^ of October 2015. The two study groups for this research project consisted of inward patients admitted for delivery to ward 3 from 1^st^ April 2015 to 1^st^ October 2015. They were between the ages of 20 to 35 years and regularly followed up at antenatal clinics at CSHW. The first group (cases) consisted of patients diagnosed with GH/PEC while the second group consisted of patients with similar ages and parity but without GH/PEC. Patients without a regular clinic follow up, without existing records or written documents regarding past pregnancies or antenatal records of this pregnancy were not recruited to the study.

Patients who had adverse pregnancy outcomes (preterm labour, PPROM, placenta previa, placental abruption or IUGR), patients with multiple pregnancies, congenital uterine abnormalities or large fibroids distorting the normal uterine architecture were also excluded from the study. All the subjects along with controls were admitted to the ward during the study period and were recruited to the study till the stipulated sample size was achieved. Retrospective analysis of data related to the GH/PEC and TM was done by using antenatal records and other available medical records. Data related to the current pregnancy was taken from bed head tickets and antenatal records which included the mother’s basic information. Descriptive statistics were calculated with percentages and 95% confidence intervals. Odds ratios and 95% confidence intervals were calculated and significances were calculated at the 5% level. Informed written consent was obtained from each patient prior to the recruitment. Confidentiality of data was maintained and would be accessible only to the investigators. Ethical approval was obtained from the ethical review committee, Faculty of Medicine, University of Colombo.

## Results

245 patients with GH/PEC (cases) and 245 patients without GH/PEC (age and parity matched) (controls) were included in the study.

As observed in table 1 below, the age and parity distribution are equal in both study groups.

**Table 1:**
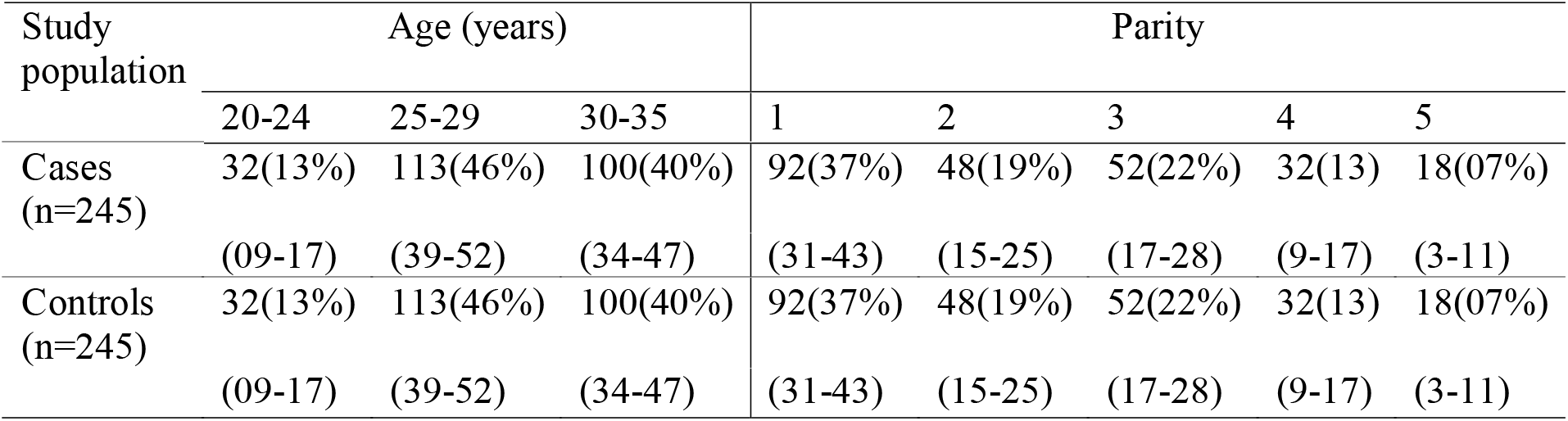
Age and Parity distribution among cases and controls (N=490)

GH and PEC can be further divided according to their severity.

Table 2 summarizes the distribution of GH/PEC among the cases according to the severity.

**Table 2:** Distribution of GH and PEC according to the severity of Hypertension.

| Category | Severity frequency (%) |  |  |
| --- | --- | --- | --- |
|  | Mild | Moderate | Severe |
| **GH** (138) | 77 (55%) | 37 (24%) | 14 (8%) |
| 95% CI | 47-63 | 20-34 | 6-16 |
| **PEC** (107) | 62 (57%) | 33 (30%) | 12 (11%) |
| 95% CI | 48-66 | 22-40 | 8-20 |

\*No cases of eclampsia

There were 56.3% cases of GH and 43.7% cases of PEC and no eclampsia patients among the cases.

In the study population, 25 patients were detected with TM in their current pregnancy. Sixteen of them had hypertension and 9 has not had hypertension (GH/PEC).

We can retrospectively tell that of the patients who had TM, 64% had subsequently developed GH or PEC.

The onset of hypertension (GH/PEC) can be early (20-32 weeks), late (after 36 weeks) or in between (32-36 weeks). According to the severity, the hypertension (GH/PEC) can be further categorized in to mild, moderate or severe. Table 4 compares the onset and severity of hypertension among the patients who had and not had a history of TM.

**Table 3.** Development of Hypertension in threatened miscarriage.

| **Study population** | **Threatened Miscarriages** | **95%CI** |
| --- | --- | --- |
| With HTN | 16 (64%) | 44-79 |
| Without HTN | 09 (32%) | 19-68 |

**Table 4.** Summary of the onset and severity of hypertension (GH/PEC) among the patients who had and not had a history of TM.

| **History (TM)** | **HTN onset in weeks** |  |  | **Severity** |  |  |
| --- | --- | --- | --- | --- | --- | --- |
|  | 20-32 | 32-36 | >36 | Mild | Moderate | Severe |
| Yes (N=16) | 10(62%) | 05(31%) | 01(06%) | 03(18%) | 06(37%) | 07(43%) |
| 95% CI | (38-81) | ((14-55) | (01-28) | (06-43) | (18-61) | (23-66) |
| No (N=229) | 29(12%) | 52(21%) | 148(64%) | 146(65%) | 64(28%) | 19(18%) |
| 95% CI | (18-17) | (01-28) | (58-70) | (57-69) | (22-34) | (05-12) |

## Discussion

Review of literature shows inconsistent evidence for association between a TM and GH/PEC. A systematic review found, incidence of GH was not significantly associated with bleeding in early pregnancy.(10) But a large prospective multicenter cohort study revealed TM with light bleeding increases the risk of PEC.(11) The current study revealed similar results which showed higher risk of PEC (OR 3.31, 95% CI 1.35-8.11) and early onset (20-32 weeks of gestation) of GH or PEC.

Also there is a significant risk of developing severe GH/PEC in the patients who had a history of TM (OR 8.59, 95% CI 2.87-25.66). It’s important to note that association of degree of bleeding during a miscarriage and risk of GH/PEC was not assessed in the current study.

Adequate oxygen and nutrient supply is an important factor that determines proper embryo development and efficient blood transport depends on the adaptive capacity of the uterine vascular system. The spiral arteries constitute the terminal portions of the uterine vasculature and penetrate the implantation and placental site.(12) Suitable implantation is dependent upon the depth and adequate amount of blood that bathes the trophoblast and thus are important factors for the proper development of the embryo. (13) Defective trophoblast invasion which leads to inadequate oxygen and nutrient transfer is a proposed pathophysiological mechanism for TM. Similar pathophysiological mechanisms are observed in PEC. Dysregulated utero-placental perfusion and placental oxidative stress, stimulate the secretion of proteins that disturb the maternal angiogenic balance. These are considered to be pre-eclampsia biomarkers.(14) Therefore, shared pathophysiological mechanisms of GH/PEC and TM may explain the observed association between these obstetric complications.

Considering the Bradford Hill criteria for causal relationship, (15) temporality and a biological gradient was observed between GH/PEC and TM. Early onset severe GH/PEC in cases with TM favors temporality and a biological gradient. It is also important to note that despite this, inconsistent conclusions were made during previous studies. Lack of multiple studies favoring a consistent positive relationship between TM and GH/PEC therefore does not favor a causal relationship.

Low dose aspirin is a well-known prophylactic treatment for PEC in high risk pregnancies and is effective when given prior to 16 weeks of gestation. Assuming published descriptive studies and systematic reviews are up to date, there is no good quality evidence available that show that low dose aspirin in TM help to prevent GH. Despite this application of best available evidence should be considered for clinical practice. Therefore, vigilant assessment of patients with TM for features of GH, PEC and EC during routine follow up visits may enable early detection and treatment. In order to go ahead and make recommendations for aspirin prophylaxis for patients with TM, it requires more consistent findings of multiple good quality randomized controlled trials. Controlled trials to test the effects of aspirin in patients with threatened miscarriage may reveal a possible preventive role of aspirin in TM.

## Conclusions

Shared pathophysiological mechanisms of GH/PEC and TM may explain the observed association between these two obstetric complications. Early onset, severe GH/PEC in cases with TM suggests a temporality and biological gradient which favors causality. Controlled trials to test the effects of aspirin in patients with threatened miscarriage may reveal a possible preventive role of aspirin in TM.

## Data Availability

Data available on request

## Notes

### Competing Interest Statement

The authors have declared no competing interest.

### Clinical Trial

Not a clinical trial

### Funding Statement

No funding

### Author Declarations

Ethical Clearance committee, Faculty of Medicine, University of Colombo ERC no; EC 15-020

